# Shared Genetic Architecture and Causal Relationship Between Diabetes, Glycemic Traits, and Cerebral Small Vessel Disease

**DOI:** 10.64898/2026.04.16.26351065

**Authors:** Keon-Joo Lee, Joo-Yeon Lee, Soo Ji Lee, Hee-Joon Bae, Joohon Sung

## Abstract

**Background:** Type 2 diabetes mellitus (T2DM) has long been considered a risk factor for cerebral small vessel disease (cSVD), yet the exact relationship between glycemic markers and cSVD remains unclear. This study explores the genetic overlap and causal associations between T2DM, glycemic indices, and cSVD phenotypes using genome-wide association studies (GWAS).

**Methods:** Using large consortium-based GWAS data, we examined relationships between T2DM, glycemic indicators (glycated hemoglobin, fasting glucose, 2-hour glucose after oral challenge, and fasting insulin), and cSVD phenotypes (white matter hyperintensity volume, lacunar stroke, cerebral microbleeds, and enlarged perivascular spaces). Our multi-level genomic strategy included: 1) identifying pleiotropic single nucleotide polymorphisms (SNPs) through PLEIO and eQTL analysis, 2) assessing genome-wide genetic correlations using LDSC and GNOVA, and 3) determining causal relationships with two-sample and multivariable Mendelian randomization analyses.

**Results:** We identified 14 pleiotropic SNPs with significant shared associations among T2DM, glycemic indicators, and cSVD phenotypes. Notably, *MICB* gene expression was elevated in brain, vascular, and pancreatic tissues, while three *HLA* genes (*HLA-DQA1, HLA-DRB1* and *HLA-DRB5*) showed reduced expression. Genetic correlation analysis revealed positive correlations between T2DM, fasting glucose, and postprandial glucose with multiple cSVD phenotypes including WMH, lacunar stroke, and perivascular spaces. Mendelian randomization demonstrated that T2DM, 2-hour glucose, and HbA1c level causally increased lacunar stroke risk (OR 1.16 [1.09-1.23], OR 1.46 [1.20-1.77], OR 1.52 [1.04-2.23], respectively). Multivariable Mendelian randomization analysis confirmed that T2DM and postprandial glucose maintained a robust direct effect on lacunar stroke independent of other cSVD phenotypes, while HbA1c did not retain significance after conditioning on cSVD imaging markers.

**Conclusions:** Our multi-level genomic analysis reveals links between T2DM, glycemic traits, and cSVD through specific genetic variants, genome-wide correlations, and causal relationships. The involvement of immune-related genes suggests potential biological mechanisms. The causal effect of postprandial glucose on lacunar stroke suggests that impaired glucose tolerance may be a relevant therapeutic target for lacunar stroke prevention.

## Introduction

Type 2 diabetes mellitus (T2DM) is a prevalent metabolic disorder that causes widespread microvascular damage across multiple organ systems, including the retina, kidneys, peripheral nerves, and increasingly recognized, the brain.^1^ Among these target organ injuries, cerebral small vessel disease (cSVD) has emerged as a critical yet incompletely understood microvascular complication of T2DM, encompassing white matter hyperintensities (WMH), lacunar infarcts, cerebral microbleeds, and enlarged perivascular spaces.^2,3^ cSVD is increasingly recognized for its importance as a well-known indicator of vascular brain health and significant contributor to stroke and cognitive decline in the aging population.^2,4,5^

The relationship between T2DM and cSVD has been investigated in several observational studies, with inconsistent findings. Some studies, such as the LADIS (Leukoaraiosis and Disability) study, have reported that subjects with diabetes show a higher rate of WMH progression and incident lacunes over a two-year follow-up period.^6^ In non-diabetic populations aged 78-79 years, glycated hemoglobin (HbA1c) has been associated with periventricular and deep WMHs.^7^ However, other studies, including the PROSPER (Prospective Study of Pravastatin in the Elderly at Risk) study, which followed participants for three years, failed to demonstrate an association between diabetes and WMH or cerebral infarcts.^8^ These conflicting results highlight the need for more robust methodological approaches to clarify whether these associations reflect true causal relationships or are confounded by other factors.

The mechanistic pathways linking T2DM and cSVD, as well as the specific glycemic indicators most relevant to cerebrovascular pathology, remain incompletely understood. Emerging genetic approaches offer new opportunities to address these limitations by examining the biological connections at multiple levels. Genome-wide association studies (GWAS) have identified numerous loci associated with T2DM and cSVD phenotypes independently.^9–14^ However, several fundamental questions remain: whether specific genetic variants exhibit pleiotropic effects across both metabolic and cerebrovascular traits, the extent of shared genetic architecture between these conditions at the genome-wide level, and ultimately, whether these genetic associations translate into causal relationships that could identify potential targets for intervention. Addressing these questions requires a comprehensive, multi-level approach that progresses from identifying specific shared genetic variants to assessing broader patterns of genetic overlap and establishing potential causal effects while minimizing confounding.

In this study, we employed a systematic, multi-level genomic approach to investigate the relationship between T2DM, glycemic traits, and cSVD phenotypes. Our analytical framework progressed logically from (1) variant-level analysis to identify specific pleiotropic genetic loci followed by colocalization analysis, to (2) genome-wide genetic correlation analysis to assess broader patterns of shared genetic architecture, and finally to (3) causal inference using Mendelian randomization (MR) techniques to determine potential causal relationships. This comprehensive approach allowed us to establish a robust body of evidence linking these conditions at both genetic and causative levels, providing insights into their shared biological mechanisms and potential clinical implications.

## Methods

### GWAS Data Sources

We utilized summary statistics from large consortium-based GWAS, restricting all analyses to individuals of European ancestry to minimize population stratification bias. All GWAS summary statistics analyzed in this study are publicly available from the original consortia. Sources and accession details for each dataset are provided in Supplementary Table S1.

For exposures, T2DM data were obtained from the DIAMANTE consortium (74,124 cases and 824,006 controls).^9^ Glycemic traits were sourced from the MAGIC consortium: fasting blood glucose (n = 200,622), HbA1c (n = 146,806), 2-hour glucose after oral glucose challenge (n = 63,396), and fasting insulin (n = 151,013).^10^

For cSVD phenotypes, we included WMH volume from UK Biobank and CHARGE consortia (n = 50,970),^11^ lacunar stroke from the International Stroke Genetics Consortium (7,338 cases and 254,798 controls),^12^ cerebral microbleeds from ADNI (3,556 cases and 22,306 controls, 94.2% European ancestry),^13^ and perivascular spaces (PVS) in white matter (WM), basal ganglia (BG), and hippocampal regions (9,324, 8,913, and 9,223 cases with 29,274, 29,990, and 29,648 controls, respectively).^14^

As this study utilized publicly available summary statistics from GWAS, no individual-level data were accessed or analyzed in this study. All contributing GWAS obtained appropriate ethical approvals and participant consent for genetic studies and data sharing. As this research involved only the secondary analysis of anonymized, publicly available summary statistics and did not require access to individual-level data or identifiable information, it was exempt from additional institutional review board approval.

### Analysis Strategy

We employed a systematic, multi-level genomic approach comprising three complementary analytical methods:

1. **Variant-level pleiotropy and colocalization analysis**: To identify specific shared genetic loci
2. **Genome-wide genetic correlation analysis**: To assess broader patterns of shared genetic architecture
3. **Causal inference analysis**: To determine potential causal relationships

This progression allowed us to build a comprehensive understanding of the genetic and causal connections between T2DM, glycemic traits, and cSVD phenotypes.

### Pleiotropic SNP Discovery and Colocalization Analysis

We employed PLEIO (Pleiotropic Locus Exploration and Interpretation using Optimal test), a summary-statistic-based framework, to map and interpret pleiotropic loci in a joint analysis of T2DM, glycemic traits, and cSVD phenotypes.^15^ This method maximizes power by incorporating genetic correlations and heritabilities of the traits into the association statistics. Our approach first identified pleiotropic SNPs (P < 5×10^-8^ from the pleiotropy analysis) that also showed associations (P < 1×10^-5^) with both glycemic traits (including T2DM) and cSVD phenotypes. We then defined independent loci by applying LD pruning (r^2^ < 0.1 using the 1000 Genomes European reference panel). Subsequently, we conducted colocalization analysis using ‘coloc’ within ± 500 kb of each top pleiotropic SNP to calculate posterior probabilities for shared causal variants between traits.^16^ We considered strong evidence of colocalization when the posterior probability for a shared causal variant (PP4) exceeded 0.8, consistent with commonly applied thresholds.

For identified pleiotropic loci, we visualized tissue-specific mRNA expression patterns based on expression quantitative trait loci (eQTL) analysis from the GTEx consortium (https://www.gtexportal.org/), focusing on brain, vascular, blood, and pancreatic tissues.^17^

### Genome-wide Genetic Correlation Analysis

We conducted genome-wide genetic correlation analyses using linkage disequilibrium score regression (LDSC) and GeNetic cOVariance Analyzer (GNOVA) to assess the shared genetic architecture between T2DM, glycemic traits, and cSVD phenotypes. LDSC estimates genetic correlation between two phenotypes by regressing the product of Z-scores from two GWAS against the LD score of each SNP, utilizing genetic covariance and LD structure information.^18,19^ GNOVA extends this approach by partitioning genetic covariance across the genome with improved statistical power, providing additional insights into local patterns of shared genetic etiology.^20^ For both methods, we used LD scores calculated from European ancestry populations in the 1000 Genomes Project as the reference panel to ensure consistency with our GWAS data sources.^21^

### Two-Sample Mendelian Randomization Analysis

To investigate potential causal relationships, we performed two-sample Mendelian randomization analyses.^21^ Instrumental variable selection involved LD clumping of SNPs with P < 5×10^-8^, using European samples from the 1000 Genomes Project as reference. Within each LD block (R^2^ < 0.001 within 10,000 kbp), the most strongly associated SNP was selected.

To reduce the influence of outlying instrumental variables and potential horizontal pleiotropy, we first applied Radial MR to identify and exclude SNPs contributing disproportionately to heterogeneity.^22^ Instrumental variables that significantly contributed to global heterogeneity (Cochran’s Q test, p < 0.05) were defined as outliers and excluded. All subsequent MR analyses were performed using this refined instrument set. Our primary analysis employed the inverse-variance weighted (IVW) method, which provides a weighted average of the causal estimates from each SNP.^23^ While the IVW approach offers the greatest statistical power, it requires all genetic variants to be valid instruments or assumes balanced pleiotropy. To address potential violations of MR assumptions, we implemented a series of complementary sensitivity analyses.^21^ The weighted median method provides consistent estimates under the "majority valid" assumption, meaning that valid instruments contribute at least 50% of the weight in the analysis.^24^ Mode-based estimation operates under the "plurality valid" assumption, identifying the most common causal effect estimate among instrument-specific estimates.^25^ MR-RAPS (Robust Adjusted Profile Score) maintains the InSIDE (Instrument Strength Independent of Direct Effect) assumption except for outliers, effectively downweighting outlying variants while accounting for weak instruments and balanced pleiotropy.^26^ MR-Egger regression operates under the InSIDE assumption and allows for unbalanced pleiotropy by introducing an intercept term that captures the average pleiotropic effect.^27^ To formally assess potential violations of MR assumptions, we examined the MR-Egger intercept for evidence of unbalanced horizontal pleiotropy and Cochran’s Q statistic for heterogeneity among instrumental variables.

### Multivariable Mendelian Randomization Analysis

To further explore whether the associations of T2DM and glycemic traits with lacunar stroke were independent of genetically predicted cSVD imaging phenotypes, we conducted multivariable Mendelian randomization (MVMR) analyses.^28^ MVMR extends the traditional MR framework by jointly modeling multiple exposures and estimating the direct effect of each exposure on an outcome conditional on the others included in the model. We performed exploratory MVMR analyses with lacunar stroke as the outcome, fitting separate models for each glycemic exposure of interest (T2DM, 2-hour postprandial glucose, and HbA1c). In each model, the glycemic exposure was jointly included with cSVD imaging phenotypes (WMH volume, cerebral microbleeds, and regional PVS phenotypes) as co-exposures to estimate conditional direct effects. These analyses were intended to explore pathway independence rather than to provide formal mediation testing. The Mendelian randomization analyses were designed, conducted, and reported in accordance with the Strengthening the Reporting of Observational Studies in Epidemiology using Mendelian Randomization (STROBE-MR) guidelines. The completed STROBE-MR checklist is provided in the Supplementary Material.

Pleiotropic locus analysis was conducted using PLEIO (https://github.com/cuelee/pleio) and colocalization analysis using coloc (v6, https://chr1swallace.github.io/coloc/). Genetic correlation was estimated using LDSC (v1.0.1, https://github.com/bulik/ldsc) and GNOVA (https://github.com/xtonyjiang/GNOVA). Two-sample Mendelian randomization was performed using the TwoSampleMR (v 0.6.22, https://mrcieu.github.io/TwoSampleMR/). Multivariable Mendelian randomization was conducted using the MVMR R package (https://github.com/WSpiller/MVMR). Statistical significance was defined as a two-sided P value < 0.05, unless otherwise specified. Given the exploratory nature of this study examining multiple related glycemic traits and cSVD phenotypes, we did not apply correction for multiple testing but instead reported 95% confidence intervals for all estimates to allow readers to evaluate the strength of evidence.

## Results

Our analytical framework progressed systematically from identifying specific pleiotropic genetic variants to assessing genome-wide genetic correlations and finally to establishing causal relationships. This multi-level approach allowed us to build a comprehensive understanding of the genetic and causal connections between T2DM, glycemic traits, and cSVD phenotypes.

### Pleiotropic SNP Discovery and Colocalization Analysis

At the variant level, our pleiotropy analysis identified 568 SNPs exhibiting significant pleiotropic associations among glycemic traits (including T2DM) and cSVD phenotypes. From these, we retained top pleiotropic SNPs representing 14 independent loci. These SNPs and their associated traits are summarized in Table 1. To further investigate whether the observed pleiotropic associations were driven by shared causal variants, we performed colocalization analyses for the identified loci. Strong evidence of colocalization (PP4 > 0.8) were found for six regions.

**Table 1.** Pleiotropic SNPs associated with T2DM, glycemic traits, and cSVD phenotypes.

| **Top**<br>**pleiotropic**<br>**SNP** | **Physical position**<br>**(GRCh37)** | **P-value**<br>**(PLEIO test)** | **Genes within**<br>**Clumping Range**<br>**($R^2 > 0.9$)** | **Associated Traits**<br>**($p < 10^{-5}$)** | **Posterior Probabilities for Shared Variant**<br>**(PP4)** |
| --- | --- | --- | --- | --- | --- |
| rs78132593 | chr1:150868102 | $4.24 \times 10^{-19}$ | *SETDB1*, *CERS2*,<br>*ANXA9* | FBG, HbA1c,<br>WMH | 99.3% (FBG & HbA1c), 87% (FBG & WMH), 81.4% (HbA1c & WMH) |
| rs2361543 | chr1:155239624 | $8.57 \times 10^{-13}$ | *SCAMP3*, *CLK2*,<br>*HCN3*, *PKLR* | HbA1c, WMH | 76.2% |
| rs2246618 | chr6:31478986 | $1.54 \times 10^{-11}$ | *MICB* | T2DM, WMH | 2.74% |
| rs532965 | chr6:32577973 | $4.62 \times 10^{-35}$ | *HLA-DRB5*, *HLA-DRB1*, *HLA-DQA1* | T2DM, HbA1c,<br>WM_PVS | 100% (T2DM & HbA1c), 80.8% (T2DM & WM_PVS), 75.5% (HbA1c & WM_PVS) |
| rs12542733 | chr8:8824858 | $1.95 \times 10^{-20}$ | - | T2DM, WMH | 0.00% |
| rs713286 | chr8:9171735 | $9.19 \times 10^{-35}$ | - | FBG, FI, WMH | 99.1% (FBG & FI), 0.04% (FBG & WMH), 0.03% (FI & WMH) |
| rs4841503 | chr8:11024326 | $5.10 \times 10^{-22}$ | *XKR6* | T2DM, FI, WMH | 42.1% (T2DM & FI), 55.1% (T2DM & WMH), 49.5% (FI & WMH) |
| rs6980507 | chr8:42383084 | $1.11 \times 10^{-29}$ | *SLC20A2, SMIM19* | HbA1c, WM_PVS | 91.1% |
| rs7940441 | chr11:46939027 | $2.50 \times 10^{-8}$ | *LRP4* | FBG, WM_PVS | 0.21% |
| rs7103835 | chr11:47461693 | $2.91 \times 10^{-45}$ | *SLC39A13, PSMC3, RAPSN* | T2DM, FBG, lacune | 0.68% (T2DM & FBG), 20.1% (T2DM & Lacune), 1.14% (FBG & Lacune) |
| rs10774624 | chr12:111833788 | $4.20 \times 10^{-25}$ | - | FI, HbA1c, lacune | 97.3% (FI & HbA1c), 87.2% (FI & Lacune), 97.9 % (HbA1c & Lacune) |
| rs55908633 | chr14:91881704 | $1.95 \times 10^{-17}$ | *CCDC88C* | T2DM, WMH | 0.00% |
| rs941897 | chr14:100599164 | $8.84 \times 10^{-10}$ | *EVL* | HbA1c, WMH | 12.4% |
| rs429358 | chr19:45411941 | $1.46 \times 10^{-28}$ | *APOE, APOC1* | T2DM, WMH, CMB | 99.9% (T2DM & WMH), 99.7% (T2DM & CMB), 99.6% (WMH & CMB) |

FBG, fasting blood glucose; FI, fasting insulin; WMH, white matter hyperintensity; WM\_PVS, white matter perivascular spaces; BG\_PVS, basal ganglia perivascular spaces; Hip\_PVS, hippocampal perivascular spaces; CMB, cerebral microbleeds.

To explore the tissue-specific expression profiles of genes located near identified pleiotropic SNPs (22 genes within an LD threshold of r^2^ > 0.9 around 14 pleiotropic SNPs), we generated a heatmap depicting the effect estimates across selected tissues. *MICB* expression was notably elevated in brain tissues, particularly in cerebellar hemisphere and cortex regions, as well as in vascular tissues including aorta and coronary arteries. Interestingly, *MICB* also showed moderate expression in pancreatic tissue, suggesting its involvement in glucose metabolism, consistent with its pleiotropic associations with T2DM and WMH. In contrast, *SMIM19* (associated with HbA1c and WM PVS) and three HLA genes (*HLA-DQA1, HLA-DRB1* and *HLA-DRB5*, associated with T2DM, HbA1c, and WM PVS) exhibited low expression across most tissues (Figure 1).

**Figure 1.**
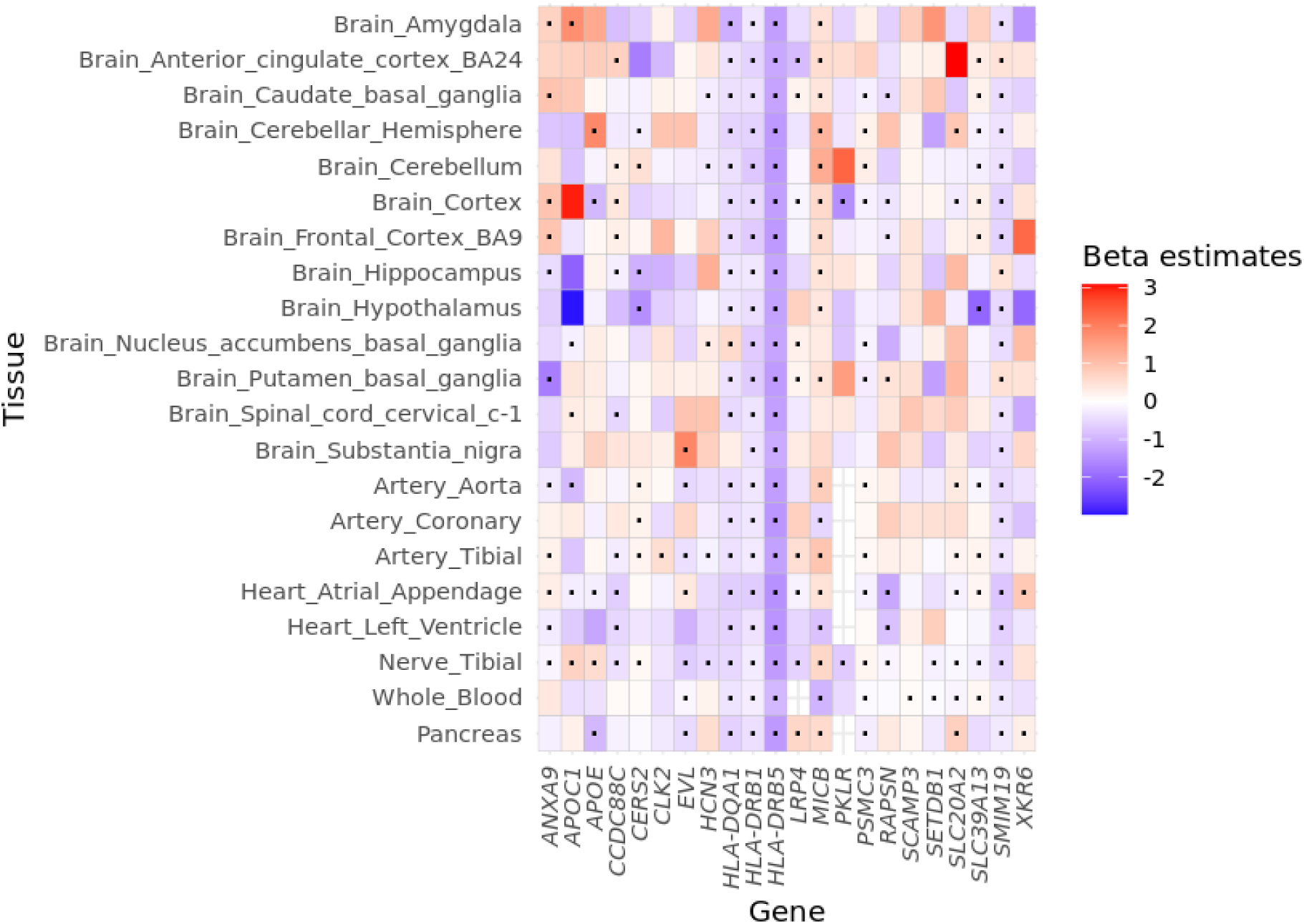
Tissue-specific expression patterns of key pleiotropic genes identified in the PLEIO analysis. The heatmap displays relative expression levels of genes across various tissues, focusing on brain, vascular, and pancreatic tissues. Red indicates elevated expression, while blue indicates reduced expression. Black dots within cells denote significant associations (FDR-adjusted q-value < 0.05).

### Genetic Correlation Analysis

Building upon our single variant findings, we next examined broader patterns of shared genetic architecture through genome-wide genetic correlation analyses using both LDSC and GNOVA methods. These complementary approaches allowed us to assess the extent of shared genetic influences between T2DM, glycemic traits, and cSVD phenotypes beyond the specific pleiotropic loci identified in our PLEIO analysis. The results are presented in Figure 2 and Supplementary Tables S2 and S3.

**Figure 2.**
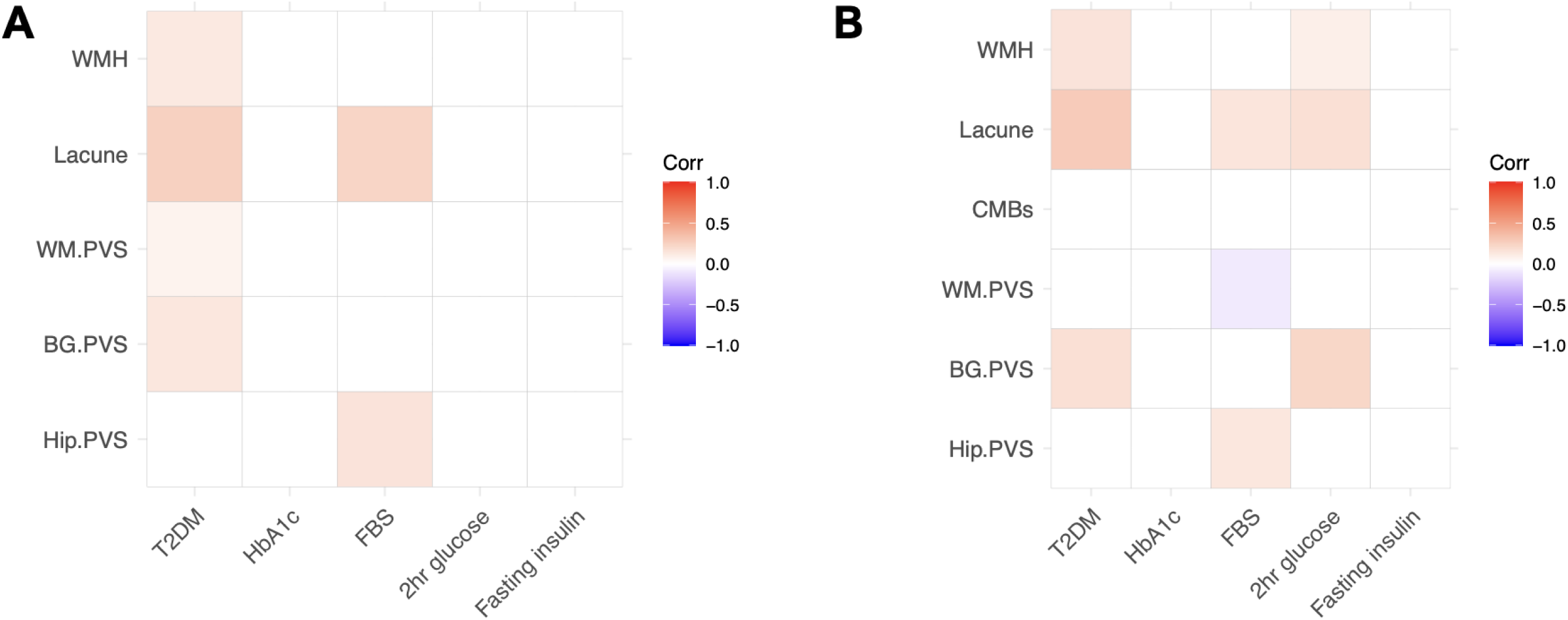
Genetic correlation analysis between T2DM, glycemic traits, and cSVD phenotypes. Heatmaps showing the statistically significant (p<0.05) genetic correlation coefficients (rg) between T2DM and glycemic traits (x-axis) and cSVD phenotypes (y-axis) using (A) LDSC and (B) GNOVA methods. Color intensity represents the strength of correlation, with red indicating positive correlation and blue indicating negative correlation. Abbreviations: WMH, white matter hyperintensity; PVS, perivascular spaces; WM, white matter; BG, basal ganglia.

The LDSC analysis revealed significant positive genetic correlations between T2DM and WMH volume (rg = 0.124, p = 0.002), lacunar stroke (rg = 0.246, p = 3.01×10⁻⁵), WM PVS (rg = 0.071, p = 0.047), and BG PVS (rg = 0.128, p = 0.025). Among glycemic traits, fasting blood glucose showed significant positive genetic correlation with lacunar stroke (rg = 0.228, p = 0.003) and hippocampal PVS (rg = 0.154, p = 0.010). Additionally, 2-hour glucose demonstrated a nominally significant correlation with WMH volume (rg = 0.124, p = 0.057). No significant genetic correlations were observed between HbA1c or fasting insulin and any cSVD phenotypes. Notably, genetic correlation estimates for cerebral microbleeds could not be reliably calculated due to insufficient statistical power.

GNOVA analyses corroborated these findings with consistent correlation patterns. The analysis revealed significant genetic correlations between T2DM and WMH volume (corr = 0.152, p = 7.44×10⁻⁶), lacunar stroke (corr = 0.277, p = 1.89×10⁻²⁰), and BG PVS (corr = 0.169, p = 0.011). Among glycemic traits, fasting blood glucose showed significant correlations with lacunar stroke (corr = 0.141, p = 0.002) and hippocampal PVS (corr = 0.134, p = 0.025), while 2-hour glucose was significantly correlated with WMH volume (corr = 0.092, p = 0.045), lacunar stroke (corr = 0.172, p = 0.002), and BG PVS (corr = 0.219, p = 0.039). Fasting blood glucose also showed a negative correlation with WM PVS (corr = - 0.090, p = 0.041). These findings suggest substantial shared genetic architecture between T2DM, glycemic traits, and specific cSVD phenotypes, with the strongest evidence for lacunar stroke and varying patterns across different cSVD imaging markers.

### Two-Sample Mendelian Randomization Analysis

Having established shared genetic architecture at both the single variant and genome-wide levels, we next investigated potential causal relationships between T2DM, glycemic traits, and cSVD phenotypes using two-sample Mendelian randomization (MR) analyses. This approach uses genetic variants as instrumental variables to infer causal effects while minimizing the influence of confounding factors. All genetic instruments demonstrated strong instrument strength (mean F-statistics range: 49.64–133.07), substantially exceeding the threshold of 10. The results are presented in Figure 3 and Supplementary Tables S4–S8.

**Figure 3.**
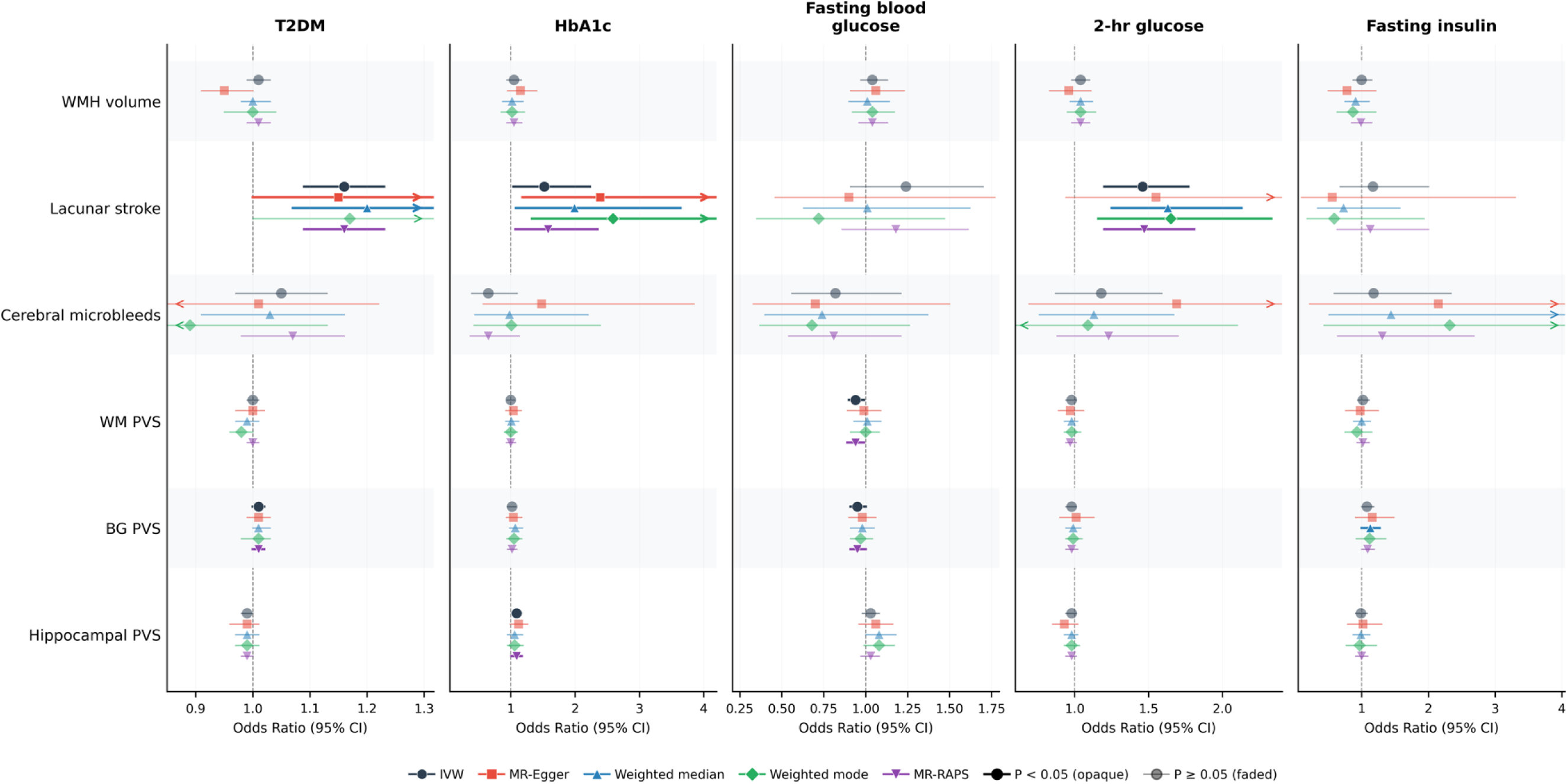
Two-sample Mendelian randomization results for glycemic traits and cSVD phenotypes. Odds ratios and 95% confidence intervals from two-sample Mendelian randomization analyses are shown for genetically predicted T2DM, HbA1c, fasting blood glucose, 2-hour glucose, and fasting insulin across cSVD phenotypes. The primary inverse-variance weighted (IVW) estimates are shown alongside sensitivity analyses including MR-Egger, weighted median, weighted mode, and MR-RAPS methods. Red indicates P < 0.05 for the IVW estimate. Abbreviations: WMH, white matter hyperintensity; PVS, perivascular spaces; WM, white matter; BG, basal ganglia.

Our primary analysis using the inverse-variance weighted (IVW) method revealed three key causal relationships. First, genetically predicted T2DM was associated with a significantly increased risk of lacunar stroke (OR = 1.16; 95% CI: 1.09–1.23; p < 0.01). This finding suggests that genetic liability to T2DM causally increases the risk of developing lacunar stroke. Higher genetically predicted 2-hour glucose levels after an oral glucose challenge (OR = 1.46; 95% CI: 1.20–1.77; p < 0.01) and HbA1c (OR = 1.52; 95% CI: 1.04–2.23; p = 0.03) were also causally linked to an elevated risk of lacunar stroke. Notably, other glycemic traits including fasting glucose, and fasting insulin did not show significant causal associations with lacunar stroke, suggesting a specific role for postprandial glucose dysregulation rather than fasting glucose level in cerebrovascular pathology.

To assess the robustness of the primary MR findings, we performed multiple sensitivity analyses. For T2DM, the weighted median (OR = 1.20; 95% CI: 1.07–1.34; p < 0.01), MR-Egger (OR = 1.15; 95% CI: 1.00–1.33; p = 0.048), and MR-RAPS (OR = 1.16; 95% CI: 1.09–1.23; p < 0.01) methods yielded directionally consistent estimates. For 2-hour glucose, the weighted median (OR = 1.63; 95% CI: 1.25–2.13; p < 0.01), weighted mode (OR = 1.65; 95% CI: 1.16–2.33; p = 0.02), and MR-RAPS (OR = 1.47; 95% CI: 1.20–1.81; p < 0.01) methods showed similar or stronger associations than the primary analysis, whereas MR-Egger produced a comparable but less precise estimate (OR = 1.55; 95% CI: 0.94–2.54; p = 0.12). For HbA1c, the weighted median (OR = 1.99; 95% CI: 1.08–3.64; p = 0.03), weighted mode (OR = 2.59; 95% CI: 1.33–5.04; p = 0.01), MR-RAPS (OR = 1.58; 95% CI: 1.07–2.35; p = 0.02), and MR-Egger (OR = 2.39; 95% CI: 1.18–4.82; p = 0.02) analyses were also directionally consistent, although estimates varied in magnitude and precision across methods. (Supplementary Tables S4 and S7).

Importantly, the MR-Egger intercept test did not indicate significant horizontal pleiotropy (p = 0.96 for T2DM, p = 0.80 for 2-hour glucose, and p = 0.14 for HbA1c), suggesting that our genetic instruments were not affecting lacunar stroke through pathways independent of the exposures. Additionally, Cochran’s Q statistic showed no significant heterogeneity among the instrumental variables for T2DM (Q = 107.86, p = 0.99), HbA1c (Q = 58.43, p = 0.74), or 2-hour glucose (Q = 6.78, p = 0.75), further supporting the validity of our MR analyses.

Interestingly, we did not observe broadly consistent causal associations across other cSVD phenotypes (e.g., WMH volume or cerebral microbleeds) despite the genetic correlations identified earlier. However, we detected selective associations for PVS phenotypes: T2DM showed a positive association with basal ganglia PVS, HbA1c showed a positive association with hippocampal PVS, and fasting blood glucose showed inverse associations with white matter and basal ganglia PVS in the IVW and MR-RAPS analyses. These findings suggest that the shared genetic architecture may not uniformly translate into direct causal effects across cSVD phenotypes and that any causal relationships may be phenotype-specific and method-dependent.

These MR findings build upon our earlier genetic discoveries by establishing potential causal relationships between T2DM, HbA1c, postprandial glucose levels, and lacunar stroke. The consistency across different MR methods and the absence of significant pleiotropy or heterogeneity strengthen the evidence for these causal associations.

### Multivariable Mendelian Randomization Analysis

As the final step in our analytical framework, we conducted multivariable Mendelian randomization (MVMR) analyses to further refine our understanding of the causal relationships. MVMR allows us to estimate the direct effect of each exposure on an outcome while accounting for the effects of other relevant exposures or potential mediators. In this case, we aimed to determine whether the effects of T2DM, 2-hour glucose levels, and HbA1c on lacunar stroke were independent of other cSVD phenotypes, which could potentially act as mediators in this relationship. The results are presented in Figure 4.

**Figure 4.**
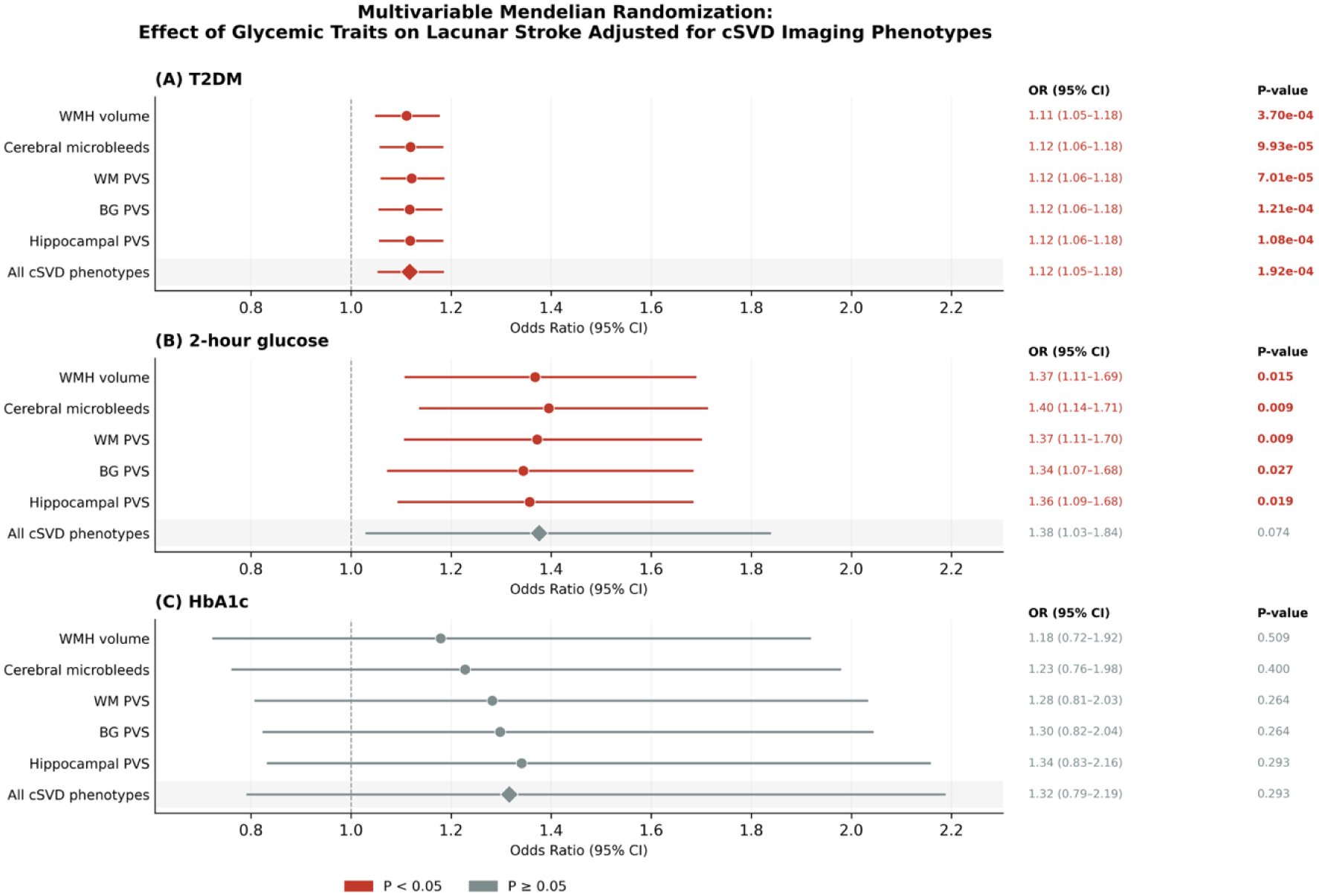
Multivariable Mendelian randomization results for the effect of T2DM, 2-hour glucose, and HbA1c on lacunar stroke. Forest plots showing odds ratios and 95% confidence intervals for the association between (A) T2DM and lacunar stroke, (B) 2-hour glucose and lacunar stroke, and (C) HbA1c and lacunar stroke in models adjusting for different cSVD phenotypes. The bottom row in each panel represents the fully adjusted model including all cSVD phenotypes simultaneously. Abbreviations: WMH, white matter hyperintensity; PVS, perivascular spaces; WM, white matter; BG, basal ganglia.

For T2DM, we conducted several MVMR models, each adjusting for a different cSVD phenotype (WMH volume, cerebral microbleeds, WM PVS, BG PVS, and hippocampal PVS) as well as a comprehensive model including all cSVD phenotypes simultaneously. Across all models, T2DM consistently maintained a significant association with lacunar stroke. When adjusting for individual cSVD phenotypes, odds ratios ranged from 1.11 to 1.12 (all p < 0.01): WMH volume-adjusted OR = 1.11 (95% CI: 1.05–1.18), cerebral microbleeds-adjusted OR = 1.12 (95% CI: 1.06–1.18), WM PVS-adjusted OR = 1.12 (95% CI: 1.06–1.18), BG PVS-adjusted OR = 1.12 (95% CI: 1.06–1.18), and hippocampal PVS-adjusted OR = 1.12 (95% CI: 1.06–1.18). In the fully adjusted model including all cSVD phenotypes simultaneously, the odds ratio was 1.12 (95% CI: 1.06–1.18; p = 1.92×10⁻⁴), demonstrating that the causal effect of T2DM on lacunar stroke persists independently of all measured cSVD phenotypes.

Similarly, for 2-hour glucose levels after an oral glucose challenge, the association with lacunar stroke remained significant across most MVMR models adjusting for individual cSVD phenotypes. Odds ratios ranged from 1.34 to 1.40: WMH volume-adjusted OR = 1.37 (95% CI: 1.11–1.69), cerebral microbleeds-adjusted OR = 1.40 (95% CI: 1.14–1.71), WM PVS-adjusted OR = 1.37 (95% CI: 1.11–1.70), BG PVS-adjusted OR = 1.34 (95% CI: 1.07–1.68), and hippocampal PVS-adjusted OR = 1.36 (95% CI: 1.10–1.68). In the fully adjusted model accounting for all cSVD phenotypes simultaneously, the point estimate remained consistent (OR = 1.38; 95% CI: 1.03–1.84) but did not reach conventional statistical significance (p = 0.074). In contrast, HbA1c did not show a significant association with lacunar stroke in any of the MVMR models, whether adjusting for individual cSVD phenotypes or in the fully adjusted model (OR = 1.32; 95% CI: 0.79–2.19; p = 0.293), suggesting that the two-sample MR signal for HbA1c may not reflect a direct, independent causal effect on lacunar stroke.

## Discussion

Our study employed a comprehensive, multi-level genomic approach to investigate the relationship between T2DM, glycemic traits, and cSVD phenotypes. By systematically progressing from single variant analysis to genome-wide correlations and finally to causal inference, we have established a robust framework of evidence linking these conditions at both the genetic and causative levels.

At the single variant level, our PLEIO analysis identified 14 pleiotropic SNPs that exhibited significant shared associations among T2DM, glycemic indicators, and cSVD phenotypes. The discovery of these specific genetic variants, particularly those near immune-related genes such as *MICB*, *HLA-DQA1, HLA-DRB1* and *HLA-DRB5* provides direct evidence of shared genetic architecture at precise genomic loci. The colocalization analyses further strengthened this evidence by demonstrating that several of these loci share causal variants influencing both metabolic and cerebrovascular traits. *MICB* encodes a stress-induced ligand that activates natural killer cells and CD8+ T cells through NKG2D receptor binding, playing a crucial role in immune surveillance of damaged cells.^29^ The elevated expression of *MICB* in brain and vascular tissues suggests a direct role in cerebrovascular pathology. Similarly, HLA-DQA1, HLA-DRB1 and HLA-DRB5, a major histocompatibility complex class II molecule, is central to antigen presentation and immune activation associated with susceptibility with infectious and autoimmune diseases.^30^ These findings implicate chronic low-grade inflammation and immune-mediated vascular injury as potential mechanisms linking glycemic dysregulation to cSVD, consistent with mechanisms proposed by previous preclinical and clinical studies.^31–33^ The convergence of metabolic and immunologic pathways may explain why standard glucose-lowering interventions have shown inconsistent effects on cerebrovascular outcomes, as they primarily target glycemic control without addressing the underlying inflammatory processes.

Building upon these single variant findings, our genome-wide genetic correlation analyses using LDSC and GNOVA revealed broader patterns of shared genetic etiology. The significant positive genetic correlations between T2DM and WMH volume, as well as between glycemic traits (particularly 2-hour glucose and fasting glucose) and lacunar stroke, indicate that the genetic overlap extends beyond the few pleiotropic loci identified in our PLEIO analysis. These genome-wide correlations suggest a more extensive shared genetic predisposition that encompasses multiple pathways and biological processes.

Having established genetic connections at both specific loci and genome-wide levels, our Mendelian randomization analyses examined causal relationships. Consistent with previous Mendelian randomization studies, we found no significant association between T2DM or glycemic traits and WMH volume, but observed robust causal effects on lacunar stroke risk.^34,35^ Notably, prior studies did not examine 2-hour postprandial glucose as a separate exposure, nor did they employ multivariable MR to disentangle direct from indirect effects or pleiotropy analysis to identify shared immune-related pathways. The specific association with 2-hour postprandial glucose is particularly noteworthy. Postprandial hyperglycemia generates acute endothelial dysfunction through excessive reactive oxygen species production, protein kinase C activation, advanced glycation end product formation, and inflammatory responses.^36,37^ Small perforating arteries supplying subcortical regions are especially vulnerable to these perturbations given their limited collateral circulation serving as an end artery.^38^ Unlike HbA1c, which reflects average glycemic control, postprandial glucose captures acute hyperglycemic excursions. Notably, this mechanism parallels the pathophysiology of coronary microvascular dysfunction in diabetes, where postprandial hyperglycemic spikes similarly induce endothelial injury in small coronary arteries, suggesting a shared microvascular vulnerability across cardiac and cerebral vascular beds.^39^ Notably, HbA1c also showed a causal association with lacunar stroke in the two-sample MR analysis, with directionally consistent and statistically significant estimates across multiple MR methods. In contrast, fasting blood glucose did not show a comparably consistent causal association with lacunar stroke in our analyses. These differential patterns suggest that both acute glycemic excursions (captured by 2-hour glucose) and chronic glycemic burden (reflected by HbA1c) contribute to lacunar stroke risk through distinct mechanisms, whereas fasting glycemia alone may be less informative for this specific cSVD phenotype.

Our multivariable Mendelian randomization analyses provided further insight into the specificity of these causal pathways. The effect of T2DM on lacunar stroke remained robust after conditioning on other cSVD imaging markers (WMH, PVS, CMB), and 2-hour glucose showed directionally consistent associations across most individual-cSVD-adjusted models. In contrast, HbA1c did not retain significance in any MVMR model, suggesting that its two-sample MR signal may reflect an indirect effect mediated through chronic structural microvascular changes—such as WMH and PVS—rather than a direct causal influence. As a marker of average glycemic exposure over the preceding 2–3 months, HbA1c likely captures cumulative vascular damage that manifests through these intermediate cSVD phenotypes, which were accounted for in the MVMR models. These differential MVMR patterns have several important implications. First, the persistent direct effect of T2DM and the attenuation of HbA1c suggest that the broader metabolic syndrome associated with diabetes influences lacunar stroke through pathways at least partially distinct from those captured by cSVD imaging phenotypes. While WMH, PVS, and CMB reflect chronic manifestations of small vessel disease visible on neuroimaging, lacunar infarcts represent acute ischemic events caused by occlusion of single penetrating arteries.^4,40^ The direct effect on lacunar stroke risk may be mediated by acute metabolic disturbances affecting endothelial function, blood-brain barrier integrity, atherosclerosis of the parent artery and thrombogenicity—processes that precede or occur independently of structural changes detectable on conventional MRI.^40–43^ Second, this suggests that current cSVD imaging markers may not fully capture the dynamic vascular dysfunction induced by glycemic dysregulation. Traditional imaging markers represent relatively stable structural changes, whereas the pathway from hyperglycemia to lacunar stroke may involve functional alterations in cerebrovascular reactivity, autoregulation, and acute thrombotic responses that are not adequately reflected in standard neuroimaging assessments.^4,44,45^

From a clinical perspective, our findings carry important implications for the management of diabetic microvascular complications beyond traditional target organs. The causal effect of postprandial glucose on lacunar stroke risk offers a potential explanation for why landmark intensive glucose control trials such as ACCORD, ADVANCE, and VADT showed inconsistent cerebrovascular benefits—these trials primarily targeted HbA1c reduction rather than postprandial glucose excursions specifically.^46–48^ Modern glucose-lowering agents with established cardiovascular benefits, including SGLT2 inhibitors and GLP-1 receptor agonists, have demonstrated vascular protective properties beyond glycemic control, including anti-inflammatory effects, improvement of endothelial function and cerebrovascular reactivity, and reduction of oxidative stress in preclinical models.^49–51^ Whether these agents confer specific protection against cSVD progression warrants investigation in dedicated clinical trials. Indeed, a recent Mendelian randomization study demonstrated that genetically predicted SGLT2 inhibition was associated with a lower risk of deep cerebral microbleeds and small vessel stroke, supporting the potential cerebrovascular benefits of these agents.^52^ While that study focused on drug-target effects, our findings complement this evidence by identifying which upstream glycemic traits—particularly postprandial glucose dysregulation—most strongly drive lacunar stroke risk. Furthermore, the identification of immune-related genetic pathways (MICB, HLA genes) linking glycemic dysregulation to cerebral microvascular disease suggests that anti-inflammatory strategies may represent promising therapeutic targets that could modulate both metabolic and cerebrovascular outcomes.

However, our study has limitations. The use of summary-level data precludes the assessment of individual-level interactions and potential confounders. Additionally, the majority of the GWAS data utilized were derived from populations of European ancestry, which may limit the generalizability of our findings to other ethnic groups. Further research in diverse populations is warranted to validate these associations across different ancestral backgrounds. Although we used GWAS from separate consortia for exposures and outcomes, partial sample overlap (e.g., UK Biobank participants contributing to both the DIAMANTE and WMH GWAS) cannot be entirely excluded, which may introduce bias toward confounded estimates. Another limitation is the potential for residual pleiotropy, despite our comprehensive sensitivity analyses.^53^ While the MR-Egger intercept test did not suggest significant horizontal pleiotropy, it is possible that some genetic variants may influence cSVD phenotypes through pathways independent of glycemic regulation. Furthermore, the relatively small number of instrumental variables available for 2-hour glucose (n = 10–13) may limit the precision of causal estimates for this exposure. However, the strong F-statistics (range: 57.46–59.83) indicate that these instruments are sufficiently robust to minimize weak instrument bias, and the consistency across multiple MR methods including MR-RAPS—which is robust to weak instruments—supports the validity of these findings. Nevertheless, replication in larger GWAS datasets for postprandial glucose traits is warranted. Finally, the cross-sectional nature of GWAS data limits our ability to infer temporal relationships and dynamic interactions between glycemic traits and cSVD progression over time.

## Conclusions

Our multi-level genomic analysis revealed important connections between T2DM, glycemic traits, and cSVD phenotypes. At the single variant level, we identified 14 pleiotropic SNPs with shared associations, particularly near immune-related genes such as *MICB* and *HLA* genes. At the genome-wide level, we demonstrated significant genetic correlations indicating broader patterns of shared genetic architecture. Our Mendelian randomization analyses provided evidence that T2DM and postprandial glucose levels have causal effects on lacunar stroke risk, with T2DM maintaining robust associations independent of other cSVD markers in multivariable analyses. While HbA1c was also causally associated with lacunar stroke in two-sample MR, its effect was attenuated in multivariable models, suggesting an indirect pathway mediated through shared cSVD mechanisms.

This progression from specific genetic variants to genome-wide correlations and causal relationships contributes to our understanding of the connections between metabolic dysregulation and cerebrovascular pathology. The involvement of immune-related genes suggests potential biological mechanisms underlying these associations. The specific link between postprandial glucose and lacunar stroke suggests that impaired glucose tolerance, as reflected by postprandial hyperglycemia, may represent a more relevant therapeutic target than fasting glycemia for lacunar stroke prevention. Given the parallel microvascular pathology shared between cerebral and coronary small vessels, these findings may also inform broader strategies for preventing diabetic microvascular complications across multiple organ systems.

## Data Availability

All GWAS summary statistics analyzed in this study are publicly available from the original consortia. Sources and accession details for each dataset are provided in Supplementary Table S1.

## Nonstandard Abbreviations and Acronyms

BG: basal ganglia
CI: confidence interval
CMB: cerebral microbleeds
cSVD: cerebral small vessel disease
eQTL: expression quantitative trait loci
FBG: fasting blood glucose
FI: fasting insulin
GNOVA: GeNetic cOVariance Analyzer
GWAS: genome-wide association study
HbA1c: glycated hemoglobin
IVW: inverse-variance weighted
LD: linkage disequilibrium
LDSC: linkage disequilibrium score regression
MR: Mendelian randomization
MR-RAPS: MR robust adjusted profile score
MVMR: multivariable Mendelian randomization
OR: odds ratio
PVS: perivascular spaces
SNP: single nucleotide polymorphism
T2DM: type 2 diabetes mellitus
WM: white matter
WMH: white matter hyperintensity

## Acknowledgments

We thank all the participants and researchers who contributed to the GWAS datasets used in this study. We acknowledge the DIAMANTE, MAGIC, UK Biobank, CHARGE, International Stroke Genetics Consortium, and ADNI consortia for making their summary statistics publicly available.

## Supplemental Material

Tables S1–S8 Figures S1–S5

